# Spatial Prevalence and Determinants of Malaria among under-five Children in Ghana

**DOI:** 10.1101/2021.03.12.21253436

**Authors:** Bedilu Alamirie Ejigu, Eshetu Wencheko

## Abstract

In Ghana malaria is an endemic disease and the incidence of malaria still accounts for 38.0% of all outpatient attendance with the most vulnerable groups being children under 5 years of age. In order to alleviate this problem, it is essential to design geographically targeted and cost-effective intervention mechanisms guided by up-to-date and reliable data and maps that show the spatial prevalence of the disease. The 2016 Ghana Malaria Indicator Survey data (N = 2,910 under-five children) were analyzed using model-based geostatistical methods with the two objectives to: (1) explore individual-, household-, and community-level determinant variables associated with malaria illness in U5 children, and (2) produce prevalence maps of malaria across the study locations in the country. The overall weighted prevalence of malaria by microscopy blood smear and rapid diagnostic tests were 20.63% (with 95% CI: 18.85% - 22.53%) and 27.82% (with 95% CI: 25.81% - 29.91%), respectively. Across regions of Ghana, the prevalence of malaria ranges from 5% in Greater Accra to 31% in Eastern region. Malaria prevalence was higher in rural areas, increased with child age, and decreased with better household wealth index and higher level of mother’s education. Given the high prevalence of childhood malaria observed in Ghana, there is an urgent need for effective and efficient public health interventions in hot spot areas. The determinant variables of malaria infection that have been identified in this study as well as the maps of parasitaemia risk could be used in malaria control program implementation to define priority intervention areas.

## 1 Introduction

Malaria is an infectious disease caused by a parasite that is transmitted from subject to subject by blood sucking female anopheles mosquitoes. Malaria is one of the major public health problems especially in Africa and Asia. In Africa, more than two-thirds of all malaria deaths occur in children under five years of age WHO (2017). Because of continual fight against malaria intervention programs, malaria infection prevalence and clinical incidence decreased by 50% and 40%, between 2000 and 2015, respectively, in Africa Bhatt et al. (2015). As presented in WHO annual report WHO (2017), there were an estimated 216 million malaria cases and 445 thousand malaria deaths in 2017. Among those cases, the majority which accounts for 91% occurred in Sub-Saharan Africa WHO (2017). Considering all malarious countries in the world, fifteen countries accounted for 80% of global malaria deaths in 2016 worldwide, and from this proportion Ghana accounts 4% WHO (2017).

In 2018, an estimated 219 million cases and 435 thousand deaths of malaria occurred world-wide, of which 200 million (92%) malaria cases were in the WHO African Region WHO (2018).

Malaria is a common disease with seasonal fluctuations in all parts of Ghana with varying degrees of prevalence across different ecological zones, and transmission occurs year-round with relatively higher prevalence in the northern part of the country. Among all outpatient visits and admissions in health facilities, 38% and 27.3%, are due to malaria cases, respectively PMI (2017); NMCP (2014). In the year 2015, among all children deaths recorded in Ghana, 48.5% are due to malaria infection. Nowadays, due to increased access to health facilities across the country, there is an increase in total outpatient department cases from year to year. Moreover, because of the accessibility of health facilities in different parts of the country, malaria testing were increased from 39% in 2013 to 78% in 2016 PMI (2017). Thus, due to such early testing and other intervention mechanisms, malaria-related mortality has declined significantly from 19% in 2010 to 4.2% in 2016 PMI (2017).

Even though, Ghana’s entire population is at risk of malaria due to different environmental and ecological factors, pregnant women and under-five children are at higher risk of severe illness due to their low immunity. Thus, interventions and preventive measures could be improved by advancing our understanding about the spatial patterns of malaria prevalence distribution and underlying determinant variables. To this effect, the first national Ghana Malaria Indicator Survey (GMIS) was conducted in 2016 with the objective to plan evidence-based intervention mechanisms. The survey dataset contains information, among others, on malaria-related burden and coverage of key intervention mechanisms in children under the age of five. Among various malaria intervention mechanisms, the use of long-lasting insecticidal nets (LLINs) is one of the main intervention mechanisms. GMIS shows that 73% of households in Ghana had at least one LLIN Ghana Statistical Service (2017). In order to minimize malaria related morbidity and mortality burden in the country by 75% by 2020, the Ghana Strategic Plan for Malaria Control aimed to increase LLINS coverage through mass campaigns and continuous distribution at antenatal clinics, child welfare clinics, and primary schools NMCP (2014).

A spatial distribution risk map of malaria is an important tool for effective planning, malaria control intervention, resource mobilization, monitoring and evaluation process in a country like Ghana where malaria is a serious health problem. To advance intervention mechanisms, spatial distribution maps of malaria prevalence across study areas have been produced at different geographical scales in African countries using geostatistical modeling approaches Adigun et al. (2015); Gosoniu et al. (2010, 2012); Riedel et al. (2010); Samadoulougou et al. (2014); Khagayi et al. (2014); Kazembe et al. (2006); Ssempiira et al. (2017) with the aim of identifying areas where greatest control effort should be focused.

Earlier maps of the geographical distribution of malaria in Ghana are available at regional, continental, and global scale Kumi-Boateng et al. (2015); Awine et al. (2017); Hay and Snow (2006); Gething et al. (2011); Bhatt et al. (2015). However, those maps may not characterize the current malaria situation in the country.

To date, to our knowledge, the only map available to depict the spatial distribution of malaria prevalence in Ghana is the one obtained from the new world malaria map Kumi-Boateng et al. (2015); Awine et al. (2017); Hay and Snow (2006); Gething et al. (2011); Bhatt et al. (2015).Evidently, that map cannot represent the current situation since it does not take into account contemporary effects of interventions and socio-economic aspects. The recent intervention strategies in PMI (2017); NMCP (2014); GHS (2017); Ghana Statistical Service (2017) that deal with malaria provide only simple descriptive statistics without addressing the spatial prevalence/distribution of malaria by considering pertinent factors related to malaria.

The current study analyzes the 2016 GMIS data using two statistical modelling approaches namely, Bayesian and likelihood-based geostatistical modelling to: (1) identify determinant variables associated with malaria risk, and (2) produce prevalence maps of malaria in under-five children across survey clusters and regions of the country. Malaria prevalence maps generated by this study would have importance in policy making to target high risk areas and to identify variables that might account for the observed spatial pattern in high risk areas.

## Methods

### Ghana MIS Data

This study is based on the 2016 GMIS, which is the first comprehensive and nationally representative malaria indicator survey conducted in Ghana, obtained from the Demographic and Health Survey (DHS) website (www.dhsprogram.com) after being granted permission. The DHS provides additional information about the 2016 Ghana MIS and sampling procedures Ghana Statistical Service (2017). The main aim of this survey was to obtain population-based estimates of malaria indicators by considering a nationally representative dataset which serves as input for strategic planning and evaluation of malaria control program Ghana Statistical Service (2017). The data were collected using computer-assisted personal interviewing system using tablet computers and paper questionnaires. Figure 1 presents the map of survey locations from which raw data were collected. The distribution of survey locations across the country was uneven - with higher concentrations of survey in more densely populated areas.

**Figure 1:**
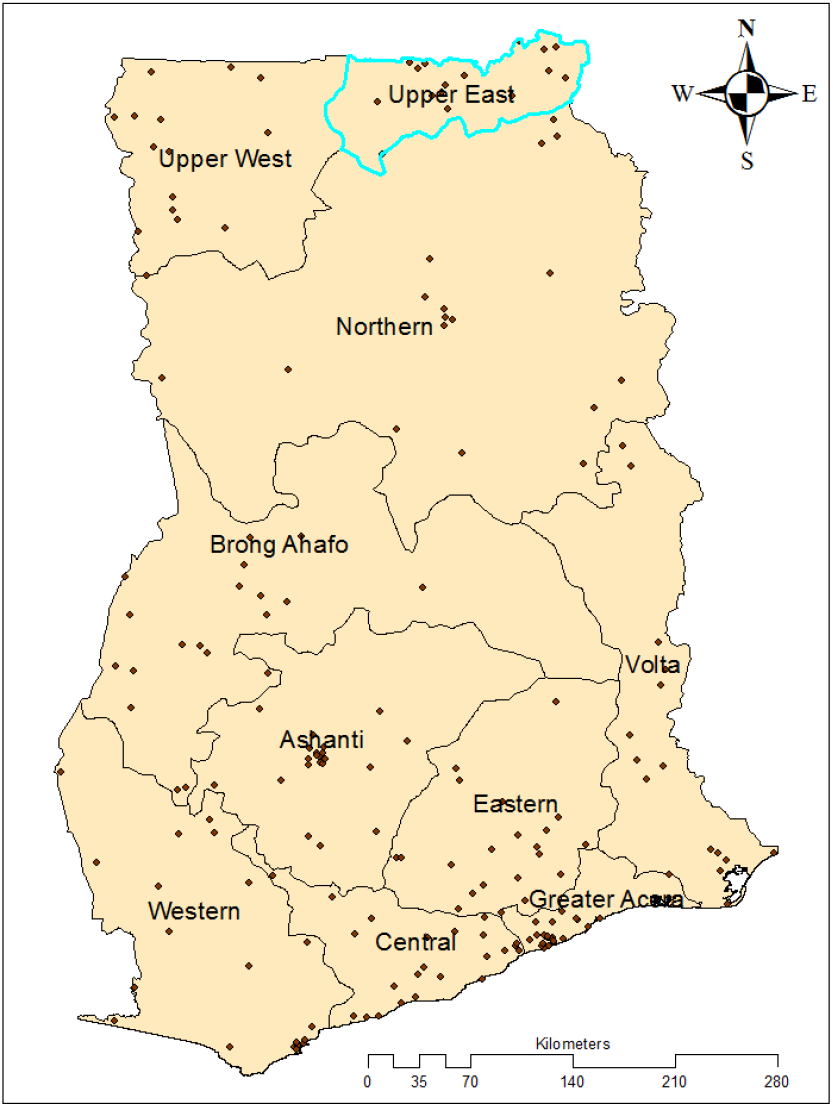
Study clusters of the 2016 GMIS. (Source: Region level country shapefiles obtained from openAfrica Code for Ghana under CC BY 4.0)

For malaria (and anaemia) testing, blood samples were collected by finger- or heel-prick from children under five years of age. After blood samples were collected, malaria tests were performed using SD BIOLINE Malaria Ag P.f./pan rapid diagnostic test (RDT) Ghana Statistical Service (2017). According to Ghana Statistical Service (2017), the tests were used to detect the histidine-rich protein II (*HRP* − *II*)^*T M*^ antigen of malaria, *Plasmodium falciparum* (P*f*), and other species in human whole blood. In addition to these tests, thick blood smears were prepared in the field and stained with Giemsa stain in order to determine the presence of Plasmodium infection at National Public Health and Reference Laboratory of Ghana. Since blood smear test results are more reliable than RDT results in identifying malaria infection, the conclusions reached will be based on blood smears test. However, this study reports the RDT results as well. The geostatistical modelling utilized individual-level variables: child age, gender, anaemia level; household-level variables: mothers education, household wealth index, and availability of bed nets in the household; and community-level variables: place of residence(urban or rural), and region.

### Statistical Analyses

Non-spatial standard statistical modelling approaches assume independence between study locations from which data are collected thereby ignoring potential spatial dependency between neighboring locations due to unobserved common exposures. To overcome that limitation, geostatistical models relate disease prevalence data with potential predictors and quantify spatial dependence via the covariance matrix structure of a Gaussian process that is facilitated by adding random effects at the observed locations Diggle et al. (1998). Such type of geostatistical models have already been used to model malaria risk at different geographical scales in different countries Adigun et al. (2015); Diggle et al. (2002); Gosoniu et al. (2012, 2010); Samadoulougou et al. (2014). In the current study the 2016 GMIS data will be analyzed by employing two modelling approaches: the likelihood-based binomial modelling and Bayesian geostatistical modelling. The following two subsections provide a brief exposure to these approaches.

### Likelihood-based Binomial Geostatistical Modelling

Let the malaria status *Y*_*ij*_ of child *i* at location *j* take the value 1 if a child has malaria, and 0 otherwise. In other words, *Y*_*ij*_ follows a Bernoulli probability distribution with *P* (*Y*_*ij*_ = 1) = *p*_*ij*_. Conditionally on a zero-mean stationary Gaussian process *S*(*l*) and additional set of study location specific random effects *Z*_*i*_, the linear predictor of the model assumes the form:

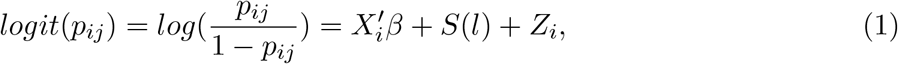

where *X*_*i*_ is a vector of child, household and community level covariates with associated regression coefficients *β*; *S* = (*S*(*l*): *l* ∈ *R*^2^) is a Gaussian process with mean zero, and variance *σ*^2^, and correlation function *ρ*(*u*) = *Corr*(*S*(*l*), *S*(*l′*)).

The model contains location specific random effects *Z*_*i*_ to account for unexplained non-spatial variation. These random effects are assumed to be independent normal distributed with a common zero mean and constant variance *τ* ^2^ (i.e. *Z*_*i*_ ∼ *N* (0, *τ* ^2^)); *τ* ^2^ is the nugget effect which accounts for the non-spatial variation. The marginal distribution of the above model is multivariate Gaussian having mean vector *Xβ* and covariance matrix Σ(*θ*) with diagonal elements *σ*^2^ + *τ* ^2^ and off-diagonal elements *σ*^2^*ρ*(*u*).

Among different parametric families of correlation functions, such as exponential, Gaussian, and spherical that have been proposed for *ρ*(*u*), Stein (1999) recommends the use of the Matern correlation function Matern (1986), given by

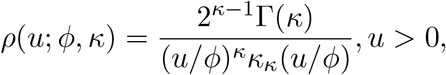

where *φ >* 0 is a scale parameter, *κ >* 0 is a shape parameter, *κ*_*κ*_() is the modified Bessel function of the second kind of order *κ*, and *u* = || *l* − *l*^*′*^ || is the Euclidean distance between two locations.

In this study the model fitting will be carried out using Monte Carlo methods by considering the PrevMap package in R Giorgi and Diggle (2017). Monte Carlo methods enable flexibility in fitting complex models and avoid asymptotic inference and the computational problems encountered in the solely likelihood-based fitting Geyer and Thompson (1992); Geyer (1994). The likelihood function for parameters *β* and *θ*^*T*^ = (*σ*^2^, *φ, τ* ^2^) is obtained by integrating out the random effects included in Equation (1).

As relates to software applications, in the computational analyses, we have used two packages. First, a weighted multilevel analysis (which takes into account survey weights) will be done using Stata StataCorp (2015). Second, the production of spatial distribution maps of malaria prevalence by survey clusters and regions Ghana, and the likelihood-based geostatistical modeling will be accomplished using R R Core Team (2017).

### Bayesian Geostatistical Modelling

The main feature of spatial data is the high dimensionality and strong dependence between the locations where data are obtained. The inferential results about geographically referenced data rely on Markov Chain Monte Carlo (MCMC) methods due to the complexity of the models Christensen et al. (2006); Christensen (2004); Diggle et al. (1998); Neal (2011). In the Bayesian modelling framework, the assumed prior distribution of parameters are combined with the likelihood function through Bayes’ theorem to obtain a posterior distribution. The current Bayesian analysis will consider prior specifications for: the range parameter *φ*, variance *σ*^2^, the nugget *τ* ^2^ and covariate parameters *β*: *φ* ∼ *Uniform*(0, 10), *log*(*σ*^2^) ∼ *N* (·; 1, 25), *log*(*τ* ^2^) ∼ *N* (·; −3, 1), *log*(*β/σ*^2^) ∼ *N* (·; 0, *σ*^2^10000).

## Results

A total of 3,078 U5 children were tested for malaria in 200 nationally representative survey clusters. Table 1 presents the overall classification of U5 children as having malaria or not based on microscopic (blood smear test) and RDT by background characteristics. The mean age of children was 32 months; 62.28% of all children lived in rural areas. The overall weighted prevalence of malaria by microscopy blood smear and RDT were 20.63% (with 95% CI:18.85% - 22.53%) and 27.82% (with 95% CI:25.81% - 29.91%), respectively. Prevalence of malaria was 28% (with 95% CI: 26% - 31%) in rural areas, and 11% (95% CI:9%-14%) in urban areas. Based on the microscopy test results, prevalence of malaria in households with and without having household mosquito nets were similar (about 21%) with slightly different confidence intervals. In addition to the above, Table 1 provides the sampling weights that are necessary in the likelihood-based geostatistical and non-spatial multilevel logistic regression models.

**Table 1:** Proportion of U5 children classified as having malaria, according to microscopy blood smear test and RDT by different background characteristics.

| Variables | Blood Smear test |  | RDT |  |
| --- | --- | --- | --- | --- |
|  | Proportion (weighted) | 95% CI | Proportion (weighted) | 95% CI |
| **Residence** |  |  |  |  |
| Urban | 0.11 | (0.09, 0.14) | 0.13 | (0.11, 0.15) |
| Rural | 0.28 | (0.26, 0.31) | 0.40 | (0.37, 0.43) |
| **Region** |  |  |  |  |
| Western | 0.24 | (0.18, 0.30) | 0.38 | (0.32, 0.45) |
| Central | 0.30 | (0.24, 0.38) | 0.44 | (0.37, 0.52) |
| Greater Accra | 0.05 | (0.02, 0.10) | 0.05 | (0.02, 0.09) |
| Volta | 0.28 | (0.23, 0.33) | 0.37 | (0.32, 0.43) |
| Eastern | 0.31 | (0.25, 0.37) | 0.34 | (0.28, 0.41) |
| Ashanti | 0.16 | (0.12, 0.22) | 0.18 | (0.13, 0.24) |
| Brong Ahafo | 0.23 | (0.18, 0.28) | 0.30 | (0.25, 0.36) |
| Northern | 0.25 | (0.21, 0.30) | 0.39 | (0.33, 0.45) |
| Upper east | 0.14 | (0.10, 0.20) | 0.26 | (0.21, 0.31) |
| Upper west | 0.22 | (0.17, 0.27) | 0.28 | (0.22, 0.34) |
| **Mother education** |  |  |  |  |
| No education | 0.30 | (0.26, 0.34) | 0.47 | (0.42, 0.52) |
| Primary | 0.25 | (0.20, 0.31) | 0.30 | (0.25, 0.36) |
| Middle | 0.20 | (0.16, 0.24) | 0.24 | (0.20, 0.27) |
| Secondary/higher | 0.05 | (0.03, 0.07) | 0.06 | (0.04, 0.09) |
| **Child age in months** |  |  |  |  |
| 6-11 | 0.17 | (0.12, 0.24) | 0.19 | (0.14, 0.26) |
| 12-23 | 0.18 | (0.14, 0.22) | 0.23 | (0.19, 0.27) |
| 24-35 | 0.19 | (0.16, 0.23) | 0.28 | (0.24, 0.33) |
| 36 -47 | 0.22 | (0.18, 0.26) | 0.32 | (0.28, 0.37) |
| 48- 59 | 0.25 | (0.21, 0.30) | 0.32 | (0.28,0.37) |
| **Child sex** |  |  |  |  |
| Male | 0.22 | (0.19, 0.25) | 0.29 | (0.26, 0.32) |
| Female | 0.19 | (0.17, 0.22) | 0.27 | (0.24, 0.30) |
| **HH Mosquito bed net** |  |  |  |  |
| No | 0.21 | (0.17, 0.26) | 0.25 | (0.21, 0.31) |
| Yes | 0.21 | (0.19, 0.23) | 0.28 | (0.26, 0.31) |
| **Wealth index** |  |  |  |  |
| Poorest | 0.37 | (0.33, 0.41) | 0.52 | (0.47, 0.56) |
| Poor | 0.29 | (0.25, 0.34) | 0.42 | (0.37, 0.48) |
| Middle | 0.17 | (0.13, 0.22) | 0.25 | (0.20, 0.30) |
| Rich | 0.12 | (0.09, 0.17) | 0.10 | (0.08, 0.14) |
| Richest | 0.02 | (0.01, 0.04) | 0.02 | (0.01, 0.04) |
| **Anaemia level** |  |  |  |  |
| Non-anaemic | 0.09 | (0.07, 0.11) | 0.13 | (0.11, 0.15) |
| Anaemic | 0.27 | (0.25, 0.30) | 0.37 | (0.34, 0.39) |

In the spatial analysis we used data from 192 clusters for which complete geo-referenced information was available at the time of the survey. As a consequence the sample size of children tested for malaria was reduced from 3.078 to 2,910. Figure 2 presents the contemporary malaria situation in survey locations. The prevalence of malaria varies from region to region: lowest in the Greater Accra region (5%) varying from 2% to 10%, and higher in the Central (30%) and Eastern (31%) regions of the country (Table 1, Fig 2 (right panel)).

**Figure 2:**
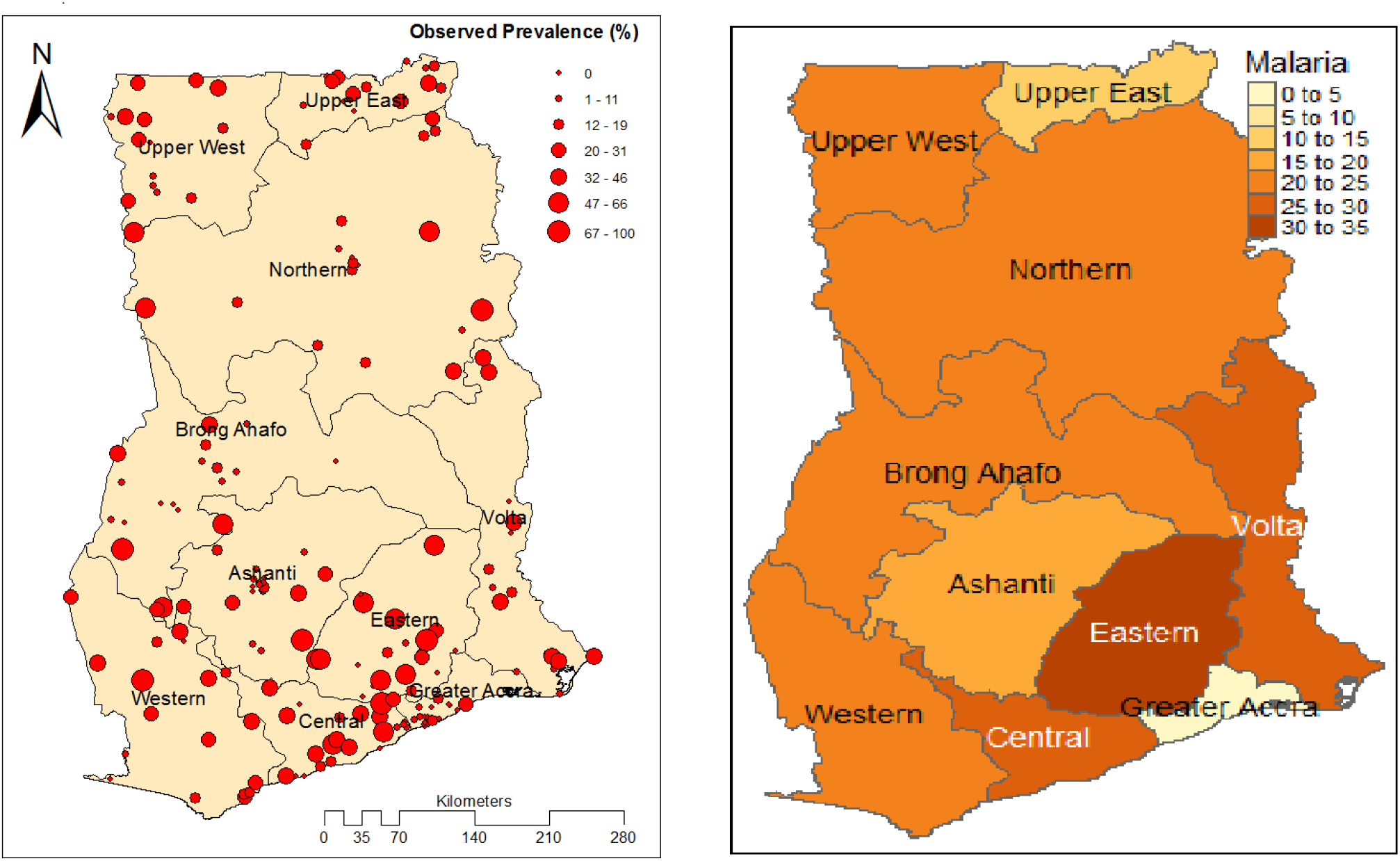
Observed malaria prevalence at survey locations (left panel) and regional estimates (right panel) in Ghana, MIS 2016. (Source: Region level country shapefiles obtained from openAfrica Code for Ghana under CC BY 4.0)

Table 2 displays the results of the analyses based on both spatial and non-spatial modellings. We note that place of residence, mothers educational level, child age, household wealth index and child anaemia level are significantly associated with malaria infection.

**Table 2:** Parameter estimates of malaria prevalence in U5 children.

| Variables | Spatial model |  |  | Non-spatial multilevel model |  |  |
| --- | --- | --- | --- | --- | --- | --- |
|  | Estimate | st.er. | P-value | Estimate | st.er. | P-value |
| **Residence**<sup>a</sup> |  |  |  |  |  |  |
| Rural | 0.87 | 0.14 | <0.0001 | 0.58 | 0.21 | 0.005 |
| **Region**<sup>b</sup> |  |  |  |  |  |  |
| Central | 0.82 | 0.25 | 0.0007 | 0.49 | 0.31 | 0.12 |
| Greater Accra | -0.64 | 0.43 | 0.135 | -0.86 | 0.37 | 0.02 |
| Volta | 0.49 | 0.27 | 0.069 | -0.35 | 0.35 | 0.31 |
| Eastern | 0.4 | 0.25 | 0.119 | 0.31 | 0.38 | 0.42 |
| Western | 0.69 | 0.25 | 0.006 | 0.12 | 0.35 | 0.76 |
| Brong Ahafo | 0.08 | 0.26 | 0.75 | -0.15 | 0.45 | 0.74 |
| Northern | 0.4 | 0.25 | 0.102 | -0.71 | 0.47 | 0.14 |
| Upper east | -0.76 | 0.26 | 0.003 | -1.18 | 0.41 | 0 |
| Upper west | 0.026 | 0.27 | 0.92 | -0.65 | 0.56 | 0.24 |
| **Mother education**<sup>c</sup> |  |  |  |  |  |  |
| Primary | -0.22 | 0.15 | 0.148 | 0.08 | 0.23 | 0.74 |
| Middle | -0.12 | 0.15 | 0.481 | 0.21 | 0.22 | 0.34 |
| Secondary/higher | -0.83 | 0.25 | 0.001 | -0.75 | 0.32 | 0.02 |
| **Child age** | 0.034 | 0.003 | <0.001 | 0.02 | 0.00 | <0.0001 |
| **Child sex**<sup>d</sup> |  |  |  |  |  |  |
| Female | -0.08 | 0.1 | 0.39 | 0.08 | 0.12 | 0.508 |
| **Household bed net**<sup>e</sup> |  |  |  |  |  |  |
| Yes | -0.16 | 0.16 | 0.344 | -0.33 | 0.2 | 0.104 |
| **Wealth index**<sup>f</sup> |  |  |  |  |  |  |
| Poor | -0.28 | 0.15 | 0.057 | -0.5 | 0.3 | 0.09 |
| Middle | -0.67 | 0.18 | 0.0001 | -1.01 | 0.26 | <0.0001 |
| Rich | -1.24 | 0.22 | < 0.0001 | -1.03 | 0.28 | <0.0001 |
| Richest | -1.96 | 0.35 | <0.0001 | -2.79 | 0.48 | <0.0001 |
| **Anaemia level**<sup>g</sup> |  |  |  |  |  |  |
| Anaemic | 1.34 | 0.12 | <0.0001 | 1.34 | 0.19 | <0.0001 |
| **Cov. Parameters** |  |  |  |  |  |  |
| $\sigma^2$ | 0.86 | 0.24 | | | | |
| $\phi$ | 0.39 | 0.78 | | | | |
| $\tau^2$ | 0.12 | 3.85 | | | | |
| **Random effect**<br>**var(cluster)** |  |  |  | 0.57 | 0.17 |  |

Ref. groups: <sup>a</sup>Urban, <sup>b</sup>Ashanti, <sup>c</sup>noeducation, <sup>d</sup>Male, <sup>e</sup>nobednet, <sup>f</sup>Poorest, <sup>g</sup>Non – anaemic

In both the spatial and non-spatial modelling approaches, anaemic children were 280% times more likely (AOR = 3.8) to be parasitaemia than non-anaemic children. In the spatial modelling, children whose mothers education is secondary or higher, were 57% less likely (AOR = 0.43) to be parasitaemia compared to children whose mothers are illiterate. The likelihood of being parasitaemia for children living in households wealth indexes that are middle and better was less likely compared with children living in households of the lowest wealth index. Children living in urban areas had a significantly low risk of having malaria compared with children in rural areas. Child sex and the availability of bed nets in the household were not significantly related with malaria prevalence in both modeling approaches. The chance of being parasitaemia for children living in rural areas was 140% higher (AOR = 2.4) than for those living in urban areas.

In the geostatistical modelling part, an exponential form was assumed for the correlation function as supported by the empirical variogram (Figure 3 (left panel)). We assessed MCMC convergence of all model parameters by examining trace plots and autocorrelation plots of the MCMC output after burn-in (Figure 3 (right panel)).

**Figure 3:**
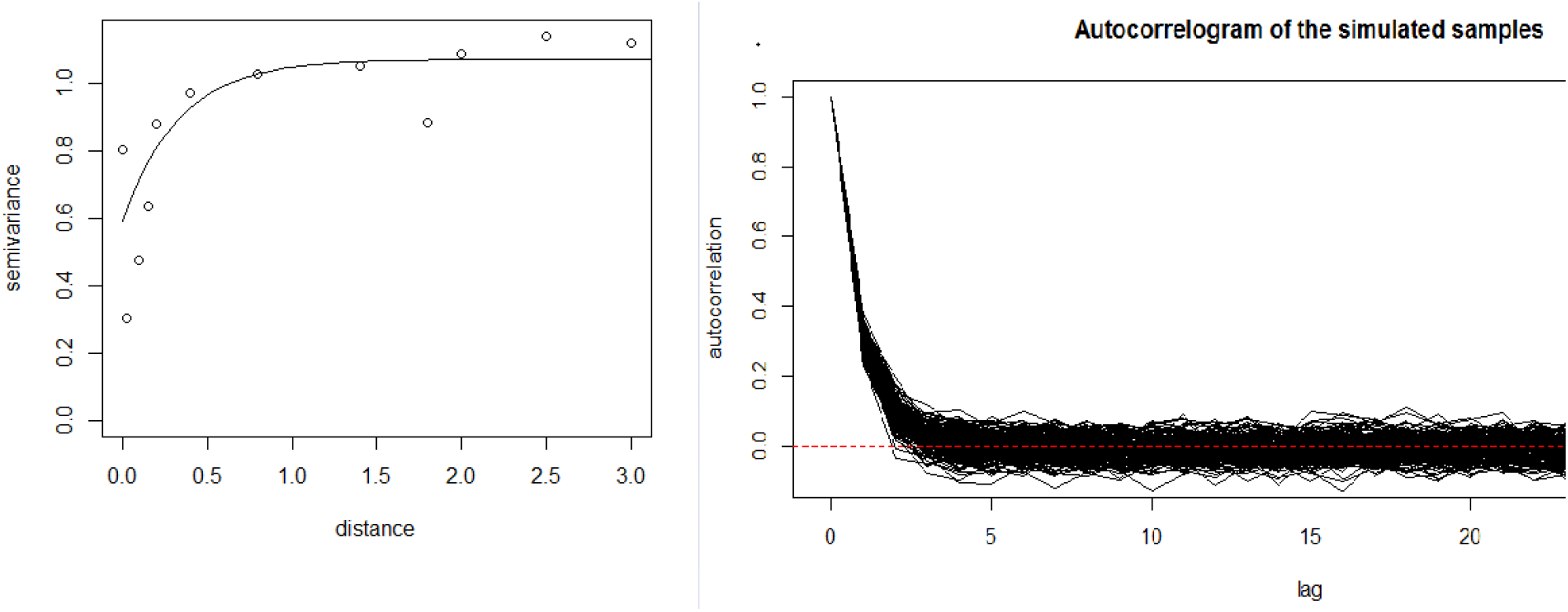
Empirical semivariogram for the empirical logit transformation of the observed prevalence with theoretical semivariogram (solid line obtained by least-squares estimation) (left panel), and autocorrelation of the simulated samples (right panel).

**Figure 4:**
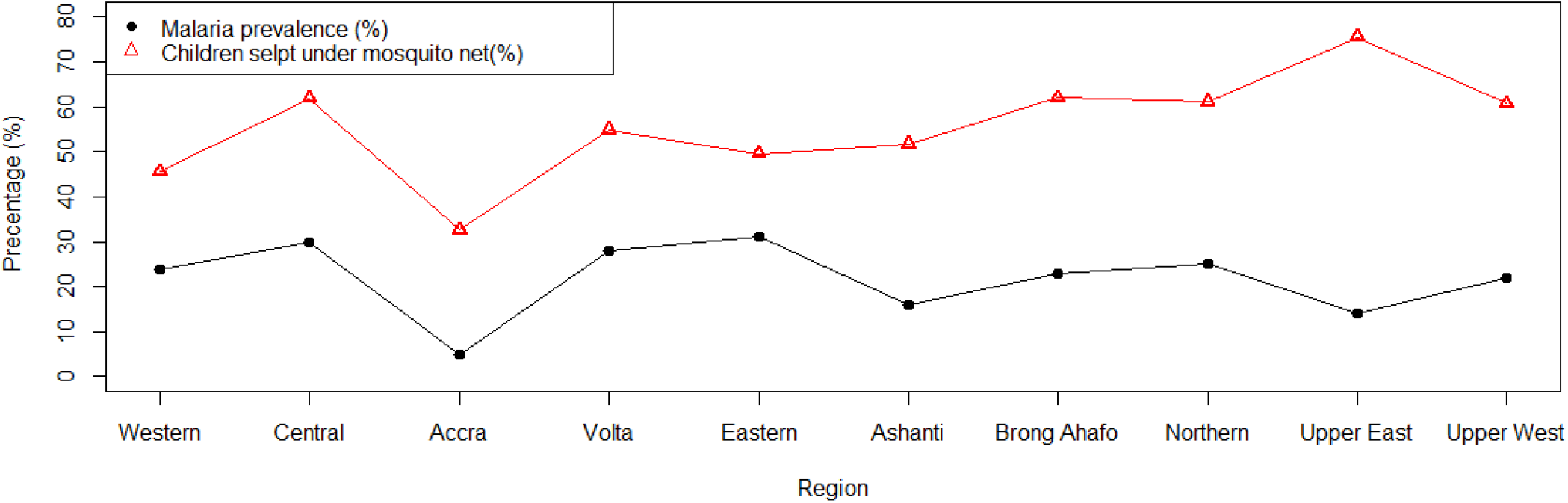
Malaria prevalence and percentage of children who slept under a mosquito net last night by region.

The non-spatial variation (*τ*^2^ = 0.12) is smaller than the spatial variation (*σ*^2^ = 0.86). In the standard non-spatial multilevel modelling, the variance of the random intercept corresponding to the cluster level variability is 0.57. Thus, employing the non-spatial model gives an intra-class level-2 correlation 0.15,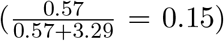. This implies that two subjects from the same cluster have a correlation of 0.15 to be parasitaemia.

In addition to the classical maximum likelihood geostatistical modelling approach, a Bayesian geostatistical model was fitted. We would like to point out that the posterior distribution for each of the corresponding regression parameters is assumed to have approximately a Gaussian distribution. Table 3 presents the posterior mean, median, standard error (st.er.) and 95% credible interval for each of the parameters considered in the Bayesian geostatistical model. The results reveal that prevalence of malaria grows with increasing age, and living in rural areas. On the other hand, bed net use, higher level of mothers education, and better household wealth index contribute to reduction of the chances of getting malaria.

**Table 3:**
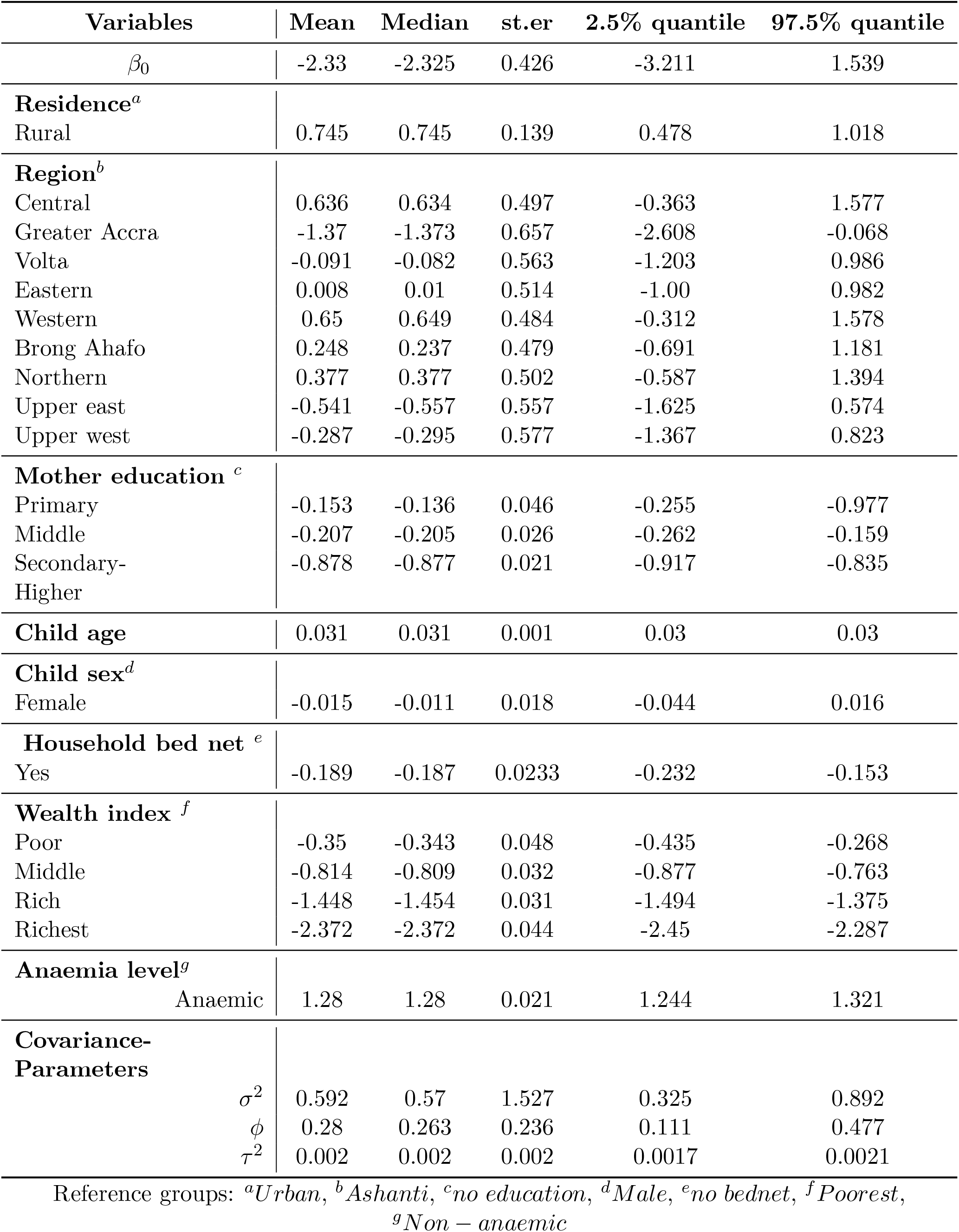
Parameter estimates of Bayesian geostatistical model.

## Discussion

This study undertook Bayesian and likelihood-based geostatistical analyses of the 2016 GMIS data with the aims to identify determinant variables of malaria risk and producing contemporary malaria risk maps of Ghana.

The current study brought to light that malaria prevalence estimates vary across different socio-demographic groups and regions of the country. The highest prevalence was observed in the Central (30% with 95% CI: 24-38%) and Eastern (31% with 95% CI: 25-37%) regions of the country. The lowest prevalence was observed in Greater Accra region (5% with 95% CI: 2-10%). Both the Bayesian and likelihood-based geostatistical analyses reveal that, children living in urban areas had significantly lower risk of having malaria compared with children in rural areas. Similar results were also found in other studies Raso et al. (2012); Gosoniu et al. (2012); Ssempiira et al. (2017). Among many other factors, malaria plays a major causative role of anaemia globally Korenromp et al. (2004); Naing et al. (2016); Adigun et al. (2015); Kassebaum et al. (2014); Ejigu et al. (2018). The overall prevalence of anaemia in under-five children in Ghana was above 75% Ewusie et al. (2014). In this study, we found that anaemic children were 3.8 times more likely to be parasitaemia than non-anemic children.

The positive association with age indicates that the older a child the higher the risk of getting infected with malaria. These findings are similar to results reported from analyses of MIS data in Nigeria Adigun et al. (2015), Angola Gosoniu et al. (2010), Tanzania Gosoniu et al. (2012), Burkina Faso Samadoulougou et al. (2014), The Gambia Diggle et al. (2002), Cote d’Ivoire Raso et al. (2012) and UgandaSsempiira et al. (2017). The studies Samadoulougou et al. (2014); Ssempiira et al. (2017); Riedel et al. (2010) have shown that the likelihood of having malaria increased as a child gets older.

Malaria prevalence among U5 Ghanian children by blood smear test was 20.63%. Evidently, this figure is much lower than prevalence rate in the four neighboring countries as depicted in Table 4. On average, 24,885 suspected malaria cases were recorded daily GHS (2017). The findings in Aryeetey et al. (2016); Gallup and Sachs (2001); Sicuri et al. (2013) call for evidence-based interventions to reduce the economic burden due to malaria and related diseases in the country. Amoah and his colleagues Amoah et al. (2017) studied the association of child growth and malaria based on DHS of 13 African countries. Their results reveal that, in all those countries, malaria had significant negative effect on child growth.

**Table 4:** Prevalence of malaria (by microscopy) in children under five years from recent surveys in neighboring countries of Ghana.

| Neighboring country | Survey year & data source | Child malaria prevalence(%) |
| --- | --- | --- |
| Burkina Faso | MIS 2014 | 29.2 |
| Cote d'Ivoire | DHS 2011-12 | 54.1 |
| Nigeria | MIS 2015 | 50 |
| Togo | DHS 2013-14 | 36 |

This study found that in rural areas, the coverage of having any mosquito bed net is 83.4%, while in urban areas the coverage is 66.1% Ghana Statistical Service (2017). Differences have also been observed across regions. Figure 5 shows that in most regions of Ghana, intervention schemes were implemented based on respective amount of burden of prevalence (higher intervention efforts were put in areas with high prevalence and vise versa). A relatively higher intervention was carried out in Upper East region.

In agreement with other studies Adigun et al. (2015); Gosoniu et al. (2010, 2012); Kazembe et al. (2006); Ssempiira et al. (2017); Diggle et al. (2002), our findings showed that malaria prevalence is strongly associated with mothers level of education, child age, wealth index and place of residence. Lower levels of household wealth indexes and mothers educational level were negatively associated with the prevalence of malaria. The findings in Gosoniu et al. (2010, 2012) and Ssempiira et al. (2017) showed a decreased risk of malaria in children from higher household wealth index and children born to mothers with higher levels of education thereby concurring with our findings.

We have produced spatially referenced malaria risk maps, the first of the kinds, for Ghana from a nationally representative geographically-referenced malaria survey data collected in a standardized way. The maps illustrate an important synopsis of prevalence of malaria in the country. Therefore, they could serve as resource in planning interventions as well as a reference in evaluating impact across administrative regions of the country. Previous maps on the spatial prevalence of malaria risk Bhatt et al. (2015); Gething et al. (2011) may not characterize the current malaria situation in the country, since those maps relied on historical survey data and that Ghana is embedded within continental and global scales. Earlier maps have led to overestimation of malaria prevalence in the country. Aside from that, they cannot reflect the current malaria prevalence (in under-five year children). We feel that the maps we have developed are a contribution to dealing with prevalence of malaria in Ghana.

## Conclusion

This study identified statistically significant determinant variables associated with malaria based on the 2016 Ghana MIS data. It also provided maps showing the contemporary spatial prevalence of malaria by survey clusters. Under-five children living in urban areas were less affected than their peers residing in rural areas. The risk of an under-five child getting malaria is higher in older children, in lower wealth index households, and among children whose mothers level of education is low. It is hoped that, household-level determinants of malaria infection that are identified and the prevalence maps developed could be of use in malaria control and intervention programs.

## Data Availability

The dataset used in this study the 2016 Ghana Malaria Indicator Survey was downloaded from the DHS program. The Dataset Terms of Use do not allow us to distribute this dataset as per data access instructions. Thus, to get access to the dataset one has to first be a registered user of the website, and download the 2016 Ghana Malaria Indicator Survey. Interested researchers will have access to the relevant data used in this study in the same way as it was accessed by the authors of this study from the aforementioned website.

## Abbreviations

AOR: Adjusted Odds Ratio
CI: Confidence Interval
DHS: Demographic and Health Survey
MCMC: Markov Chain Monte Carlo
MIS: Malaria Indicator Survey
LLINs: Long-lasting In-secticide Nets
RDT: Rapid diagnostic tests
U5: Under five years of age children
WHO: World Health Organization

